# The role of pulmonary metastasectomy (PM) for non-primary lung cancer: Umbrella review of meta-analyses

**DOI:** 10.1101/2024.04.19.24306087

**Authors:** Wongi Woo, Brandon Park, Awranoos Ahadi, Liam il-young Chung, Chan Mi Jung, Ankit Bharat, Young Kwang Chae

## Abstract

**Introduction:** Due to heterogeneous characteristics of primary cancers, the efficacy of pulmonary metastasectomy(PM) in non-primary lung cancers has not been investigated other than colorectal cancers. This study aims to investigate the clinical outcomes of PM for non-primary lung cancer.

**Methods:** A systematic search for meta-analyses on PM for non-primary lung cancers was conducted, encompassing publications up to January 3, 2024. The analysis included seven primary cancer types: renal cell, breast, adrenocortical, head and neck cancers, melanoma, germ cell tumors, and sarcoma. Primary outcomes, overall survival, and recurrence rates post-PM were assessed using random-effect models to account for study heterogeneity.

**Results:** This study included 16 systematic-review articles and 101 individual studies, involving 10,277 patients who underwent PM for non-primary lung cancer. Patients had a mean age of 48.0 years, with 68.4% being male. About half of the patients(47.1% [95%CI 40.8-53.5] presented with multiple metastatic lesions, and complete R0 resection achieved in 87.2% [95%CI 83.0-90.8]. The pooled 5-year overall survival (OS) rate post-PM was 41.2% [95%CI, 37.1-45.4%]. Patients with germ cell tumors demonstrated significantly higher survival rate than other cancers(p<0.05), while patients with melanoma exhibited the poorest outcome(p<0.05). During follow-up, 57.6%[95%CI 46.4-68.1] had recurrence; 48% of them had intrathoracic-only recurrence and 52% had extra-thoracic recurrence.

**Conclusion:** This study underscores the survival benefits associated with PM. Overall survival rates following PM do not significantly differ based on primary cancer types, except for germ cell tumors and melanoma. These findings highlight the importance of recognizing and incorporating PM into clinical practice when appropriate.

**Synopsis:** Surgical resection for pulmonary metastasis from cancers other than lung cancer still holds a significant role in improving a patient’s prognosis. Though the recurrence rate is high, it could improve the chance of survival regardless of the primary cancer types.

## Introduction

Lung is one of the organs with frequent cancer metastasis. Most patients with lung metastases are primarily treated with systematic treatments.^1,2^ These treatments are designed to control both the primary and distant metastatic lesions. Surgery alone is rarely performed in these metastatic lesions and usually combined with neoadjuvant or adjuvant treatments. However, in cases where the primary cancer is well-controlled, pulmonary metastasectomy (PM) may be pursued to achieve oncologic cure and/or survival benefits.

PM was first performed in 1939 for renal cell cancer metastasis,^3^ and pneumonectomy for extensive lung metastasis was first done in 1958.^4^ After several reports were published, its clinical benefits have been widely accepted, with PM now constituting 10.2% of all general thoracic surgeries in Japan.^5^ The number of PM procedures in the United States has also increased over time, with improvements in surgical techniques and reduced perioperative morbidity/mortality risks.^6^

In terms of primary cancer types, colorectal cancer is well-known for higher rates of pulmonary metastasis, and there have been many reports demonstrating favorable clinical outcomes following PM.^7^ Renal cell carcinoma, sarcoma, head and neck cancer, germ cell tumor, and breast cancer are also known for their frequent metastasis to lung. Compared to its high prevalence, however, the evidence for PM has been limited. Expert consensuses from thoracic surgeons have mentioned the scarcity of evidence regarding the role of PM due to the absence of randomized clinical trials and comparative survival groups, inconsistent description of local or systemic treatments, and other factors.^8^

Therefore, this study aims to analyze patients’ characteristics and clinical outcomes of PM across various primary cancer types. This will provide oncologists and surgeons with a comprehensive understanding of this procedure’s efficacy and applicability.

## Methods

This umbrella meta-analysis study was conducted according to the Preferred Reporting Items for Systematic Reviews and Meta-Analyses (PRISMA) 2020 statement (Supplementary Table S1). The study protocol was registered in PROSPERO (number: CRD42024502761).

### Search Strategy and Eligibility Criteria

Relevant studies were systematically searched in electronic databases, including PubMed/MEDLINE, EMBASE, and Scopus from their inception to January 5, 2024. The search strategy comprised of the following terms: (“pulmonary metastasis” or “lung metastasis”) AND (“cancer” or “malignancy”) AND (“surgery” or “operation”) AND (“review” or “meta-analysis”).

Eligibility criteria included (1) studies about PM from non-primary lung cancer, (2) meta-analysis studies written in English and published in a peer-reviewed journals from inception to January 5, 2024. We excluded studies with the following criteria: (1) systematic review cases without meta-analysis, (2) studies of PM results involving mixed types of primary cancer, (3) studies about PM from colorectal cancers, (4) narrative reviews, (5) studies with limited clinical data such as overall survival, and (6) studies written in languages other than English.

Three reviewers (W.W., B.P., and A.A.) independently screened papers based on title/abstract/full-text review according to the above criteria. Disagreements between authors were resolved through consultation with fourth author (Y.K.C.).

### Data Extraction

Three reviewers (W.W., B.P., and A.A.) independently extracted the relevant information, including the first author, publication year, study type and period, mean age, sex (male, %), types of surgery, clinical outcome, and articles included in each meta-analysis. The primary outcome of the quantitative meta-analysis was the overall survival rate (3-year, 5-year, and 10-year) post-PM. The secondary outcome was recurrence rate post-PM.

### Quality Assessment

The quality of each eligible systematic meta-analysis study was independently analyzed by two reviewers (B.P. and A.A.). If the evaluation was unclear, a third author (W.W.) was involved in the process. The Assessment of Multiple Systematic Reviews (AMSTAR-2) tool was used to assess the quality of the included studies. AMSTAR-2 consists of 16 items and classifies the quality level into critically low, low, moderate, and high.

### Statistical Analysis

Re-meta-analysis was performed after extracting eligible studies from each meta-analysis. First, the list of included studies in each meta-analysis was reviewed and checked whether each meets the eligibility criteria. Second, duplicates of individual studies were excluded based on author name, institution, and publication year. Third, re-meta-analysis was conducted by the type of primary cancer. Due to different designs and populations from these studies, a high degree of heterogeneity was expected. For each meta-analysis, individual studies were reanalyzed to estimate the summary effects, 95% confidence interval (CI), and p-values. The heterogeneity of individual studies was assessed using the inconsistency of the I2 metric and the p-value of the Cochrane Q test. All re-analyses in this study were performed using R version 4.0.4 (R Core Team, R Foundation for Statistical Computing, Vienna, Austria). Survival outcomes were subsequently stratified by the type of primary cancer, and their 95% CI were compared to identify any differences. All statistical tests were two-sided; p-values <0.05 were considered statistically significant.

## Results

### Study Identification

The systematic search initially yielded 612 studies after excluding duplicates, and finally 16 studies were enrolled for the final analysis.^9–24^ These 16 meta-analysis studies were categorized by seven primary cancer types, and studies involved in each meta-analysis was retrieved, consisting of 102 studies in total. Most of the included meta-analyses were assessed to have low or critically low levels by the AMSTAR-2 tool (Supplementary Table S2).

### Perioperative Patients’ Characteristics by Primary Cancer Type

Table 1 describes patients’ characteristics of the included studies according to each primary cancer type. The mean age of patients ranged from 29.6 (germ cell tumors) to 60.5 (renal tumors) years. Supplementary Figure S1 shows the 95% CIs of age by primary cancer type. Germ cell tumor, sarcoma, and adrenocortical cancer patients tended to be in a relatively younger age group than patients with other cancers.

**Table 1.** Pooled analysis of included studies with patients’ characteristics.

| Primary Cancer type | No. studies | N | Age, years | Male | Solitary | Multiple | Unilateral | Bilateral | DFI, months |
| --- | --- | --- | --- | --- | --- | --- | --- | --- | --- |
| Breast Cancer | 15 | 1373 | 54.5<br>[53.5-55.6]<br>(I<sup>2</sup>=64.1%,<br>p=0.008) | 0.15%<br>(2/1373) | 64.8%<br>[53.8, 74.4]<br>(I<sup>2</sup>=80.0%,<br>p<0.001) | 35.2%<br>[25.6, 46.2]<br>(I<sup>2</sup>=80.0%,<br>p<0.001) | 84.4%<br>[67.9; 93.2]<br>(I<sup>2</sup>=93.1%, p<0.001) | 19.7%<br>[4.5, 56.3]<br>(I<sup>2</sup>=93.1%,<br>p<0.001) | 60.0<br>[46.7, 73.3] |
|  |  | k | 14 | 15 | 11 | 11 | 3 | 3 | 4 |
| Germ cell tumor | 9 | 878 | 29.6<br>[28.2-31.1]<br>(I<sup>2</sup>=85.3%<br>p<0.0001) | All male | 29.2%<br>[24.6-34.4]<br>(I<sup>2</sup>=0.0%,<br>p=0.770) | 70.8%<br>[65.6-75.5]<br>(I<sup>2</sup>=0.0%,<br>p=0.770) | 56.3%<br>[35.5-75.2]<br>(I<sup>2</sup>=88.8%, p=0.003) | 43.7%<br>[24.8-64.6]<br>(I<sup>2</sup>=88.8%,<br>p=0.003) | 22.5<br>[18.1, 26.9]<br>(I<sup>2</sup>=48.7%<br>p=0.435) |
|  |  | k | 9 |  | 3 | 3 | 2 | 2 | 2 |
| Melanoma | 12 | 1650 | 55.4<br>[52.3, 58.5]<br>(I<sup>2</sup>=96.0%,<br>p<0.0001) | 66.1%<br>[62.1, 69.8]<br>(I<sup>2</sup>=62.9%,<br>p=0.0008) | 60.0%<br>[46.6, 72.0]<br>(I<sup>2</sup>=91.4%,<br>p<0.0001) | 40.0%<br>[28.0, 53.4]<br>(I<sup>2</sup>=91.4%,<br>p<0.0001) | 84.5%<br>[65.2, 94.1]<br>(I<sup>2</sup>=93.0%,<br>p<0.0001) | 15.5%<br>[5.9, 34.8]<br>(I<sup>2</sup>=93.0%,<br>p<0.0001) | NA |
|  |  | k | 11 | 14 | 11 | 11 | 4 | 4 |  |
| Adrenocortical carcinoma | 2 | 50 | 41.1<br>[37.4, 44.7]<br>(I<sup>2</sup>=0.0%,<br>p=0.518) | 32.2%<br>[20.7, 46.4]<br>(I<sup>2</sup>=0.0%,<br>p=0.425) | 30.1%<br>[19.0, 44.1]<br>(I<sup>2</sup>=0.0%,<br>p=0.622) | 69.9%<br>[55.9, 81.0]<br>(I<sup>2</sup>=0.0%,<br>p=0.622) | NA | NA | NA |
|  |  | k | 2 | 2 | 2 | 2 |  |  |  |
| Renal tumor | 18 | 1666 | 60.5<br>[59.3, 61.6]<br>(I<sup>2</sup>=82.7%,<br>p<0.0001) | 72.8%<br>[70.5, 75.0]<br>(I<sup>2</sup>=48.6%,<br>p=0.015) | 43.4%<br>[37.5, 49.6]<br>(I<sup>2</sup>=77.4%,<br>p<0.0001) | 56.6%<br>[50.4, 62.5]<br>(I<sup>2</sup>=77.4%,<br>p<0.0001) | 71.2%<br>[68.3, 73.9]<br>(I<sup>2</sup>=49.6%, p=0.022) | 28.8%<br>[26.1, 31.7]<br>(I<sup>2</sup>=49.6%,<br>p=0.022) | NA |
|  |  | k | 18 | 16 | 14 | 14 | 13 | 13 |  |
| Head and Neck cancers | 29 | 2353 | 54.8<br>[52.6, 57.0]<br>(I<sup>2</sup>=85%,<br>p<0.0001) | 71.7%<br>[66.4, 76.5]<br>(I<sup>2</sup>=85.9%,<br>p<0.0001) | 66.0%<br>[53.2, 76.8]<br>(I<sup>2</sup>=87.1%,<br>p<0.0001) | 24.6%<br>[15.5, 36.9]<br>(I<sup>2</sup>=86.2%,<br>p<0.0001) | 77.1%<br>[65.0, 85.9]<br>(I<sup>2</sup>=90.7%,<br>p<0.0001) | 19.2%<br>[11.9, 29.6]<br>(I<sup>2</sup>=87.7%,<br>p<0.0001) | 46.1<br>[25.0, 67.3]<br>(I<sup>2</sup>=95.6%,<br>p<0.0001) |
|  |  | k | 20 | 29 | 17 | 15 | 13 | 15 | 5 |
| Sarcoma | 16 | 2307 | 40.8<br>[31.5, 50.1]<br>(I<sup>2</sup>=99.7%,<br>p<0.0001) | 39.3%<br>[26.4, 53.9]<br>(I<sup>2</sup>=94.0%,<br>p<0.0001) | NA | NA | 43.2%<br>[25.2, 63.3]<br>(I<sup>2</sup>=94.3%,<br>p<0.0001) | 18.4%<br>[12.3, 26.6]<br>(I<sup>2</sup>=79.6%,<br>p=0.001) | NA |
|  |  | k | 10 | 15 |  |  | 4 | 5 |  |

Data is presented as value with [95% confidence interval]
DFI, disease-free interval; k, number of included studies for analysis; N, total number of participants; NA, not available.

The number of lung metastatic lesions varied among primary cancer types, with bilateral involvement being dominant in germ cell tumors (70.8%) and adrenocortical cancers (69.9%). However, most patients had unilateral involvement of pulmonary metastasis, with the highest percentage seen in melanoma (84.5%) and the lowest in germ cell tumors (56.3%). Information regarding the disease-free interval between primary cancer and pulmonary metastasis was not reported in most studies, but it ranged from 22.5 to 60.0 months in available reports.

In terms of surgical extent, most patients underwent sublobar resection, ranging from 63.8% to 89.0%, except for sarcoma (34.9%) (Table 1). However, pneumonectomy also accounted for 1.4% to 5.8% of patients. In general, 82.9% to 94.2% patients had R0 complete resection; it implies surgical challenges associated with PM due to its multiple involvements.

### Clinical Outcome by Primary Cancer

Figure 1 describes the proportion meta-analysis of OS in all included studies. The 5-year OS of all participants was 41.0% ([95% CI 37.0-44.8%], I2=89.9%, p<0.0001), and it was highest among patients with germ cell tumors (81.3% [95% CI 73.3-87.3%], I2=76.2%, p=0.0003). Figure 2 shows the summarized 5-year OS according to primary cancer. In general, most of them had similar clinical outcomes,except for melanoma, which showed the lowest rate at 25.3%, and germ cell tumors, which exhibited the highest at 81.3% (Table 2). Furthermore, 10-year OS post-PM data were available in four types of primary cancers. It ranged from 24.5% to 26.9% other than 75.2% in germ cell tumor (Table 2). Additionally, recurrence rates post-PM is depicted in Table 2, and it was high (63.2 to 74.5%) in most cancers other than germ cell tumors, which had a lower recurrence rate of 26.5% [95% CI 15.4-41.7%].

**Table 2.** Pooled analysis of metastasectomy types and clinical outcomes.

| Primary Cancer type | No. studies | N | Extent of metastasectomy |  |  | Complete resection rate | Recurrence rate | Overall survival rate |  |  |
| --- | --- | --- | --- | --- | --- | --- | --- | --- | --- | --- |
|  |  |  | Sublobar resection | Lobectomy | Pneumonectomy |  |  | 3-year | 5-year | 10-year |
| Breast Cancer | 15 | 1373 | 69.0%<br>[48.2-84.2]<br>(I<sup>2</sup>=95.0%,<br>p<0.001) | 28.2% [14.0-48.4]<br>(I<sup>2</sup>=95.0%,<br>p<0.001) | 1.9% [0.7-4.8]<br>(I<sup>2</sup>=60.6, p=0.037) | 82.7%<br>[72.0-90.0]<br>(I<sup>2</sup>=85.8%,<br>p<0.001) | 63.2%<br>[49.6-75.0]<br>(I<sup>2</sup>=90.9%,<br>p<0.001) | 61.2% [53.7-68.3]<br>(I<sup>2</sup>=65.4%,<br>p=0.005) | 44.1% [37.6-50.1]<br>(I<sup>2</sup>=84.3%,<br>p<0.0001) | 26.7% [15.8-41.5]<br>(I<sup>2</sup>=94.2%,<br>p<0.0001) |
|  |  | k | 7 | 7 | 5 | 8 | 10 | 8 | 14 | 6 |
| Germ cell tumor | 9 | 878 | 85.1%<br>[74.2, 91.9]<br>(I<sup>2</sup>=92.1%<br>p<0.0001) | 10.4%<br>[7.8, 13.6]<br>(I<sup>2</sup>=0.0%,<br>p=0.669) | 1.9% [1.0, 3.7]<br>(I<sup>2</sup>=0.0%, p=0.901) | 94.2%<br>[79.9, 98.5]<br>(I<sup>2</sup>=89.3%,<br>p<0.001) | 26.5%<br>[15.4, 41.7]<br>(I<sup>2</sup>=86.9%,<br>p<0.0001) | NA | 81.3% [73.3, 87.3]<br>(I<sup>2</sup>=76.2%,<br>p=0.001) | 75.2% [66.8, 82.0]<br>(I<sup>2</sup>=73.5%,<br>p=0.005) |
|  |  | k | 5 | 4 | 4 | 5 | 6 |  | 7 | 5 |
| Melanoma | 12 | 1650 | 70.2%<br>[60.2, 78.5]<br>(I<sup>2</sup>=84.7%,<br>p<0.0001) | 24.6% [19.3, 30.9]<br>(I<sup>2</sup>=67.3%,<br>p=0.002) | 5.2% [3.8, 7.0]<br>(I<sup>2</sup>=0.0%, p=0.767) | 83.7%<br>[76.2, 89.2]<br>(I<sup>2</sup>=81.4%,<br>p<0.0001) | 74.5%<br>[63.4, 83.2]<br>(I<sup>2</sup>=82.4%,<br>p=0.0001) | 37.0% [30.6, 43.9]<br>(I<sup>2</sup>=79.3%,<br>p<0.0001) | 25.3% [22.0, 29.0]<br>(I<sup>2</sup>=67.0%,<br>p<0.0001) | NA |
|  |  | k | 9 | 9 | 4 | 9 | 5 | 10 | 16 |  |
| Adrenocortical carcinoma | 2 | 50 | 89.0%<br>[80.2, 94.2]<br>(I<sup>2</sup>=0.0%,<br>p=0.912) | 9.8%<br>[5.0, 18.4]<br>(I<sup>2</sup>=0.0%,<br>p=0.712) | 1.80% | 83.30% | NA | 57.9% [43.9, 70.8]<br>(I<sup>2</sup>=0.0%,<br>p=0.536) | 34.6% [22.6, 49.0]<br>(I<sup>2</sup>=38.9%,<br>p=0.201) | NA |
|  |  | k | 2 | 2 | 1 | 1 |  |  | 2 |  |
| Renal tumor | 18 | 1666 | 80.5%<br>[69.5, 87.7]<br>(I<sup>2</sup>=80.7%,<br>p<0.0001) | 20.1% [14.2, 27.6]<br>(I<sup>2</sup>=93.4%,<br>p<0.0001) | 5.8% [2.1, 14.6]<br>(I<sup>2</sup>=90.0%, p<0.0001) | 87.8%<br>[81.3, 92.2]<br>(I<sup>2</sup>=80.0%,<br>p<0.0001) | 65.1%<br>[39.4, 84.3]<br>(I<sup>2</sup>=93.6%,<br>p<0.0001) | 59.7% [56.6, 62.8]<br>(I<sup>2</sup>=48.9%,<br>p=0.034) | 44.8% [39.4, 50.3]<br>(I<sup>2</sup>=77.4%,<br>p<0.0001) | 24.5% [18.4, 31.9]<br>(I<sup>2</sup>=79.0%,<br>p<0.0001) |
|  |  | k | 16 | 16 | 9 | 15 | 6 | 11 | 16 | 10 |
| Head and Neck cancers | 29 | 2353 | 63.8%<br>[38.2, 83.4]<br>(I<sup>2</sup>=95.0%,<br>p<0.0001) | 20.0% [11.5, 32.3]<br>(I<sup>2</sup>=91.3%,<br>p<0.0001) | 4.4% [1.4, 12.7]<br>(I<sup>2</sup>=77.7%, p<0.0001) | 85.9%<br>[24.7, 99.1]<br>(I<sup>2</sup>=97.9%,<br>p<0.0001) | 71.3%<br>[24.4, 95.0]<br>(I<sup>2</sup>=87.4%,<br>p=0.001) | 54.9% [36.6, 72.0]<br>(I<sup>2</sup>=87.3%,<br>p<0.0001) | 38.6% [31.6, 46.1]<br>(I<sup>2</sup>=83.1%,<br>p<0.0001) | 26.9% [11.0, 52.2]<br>(I<sup>2</sup>=90.0%,<br>p<0.0001) |
|  |  | k | 13 | 15 | 7 | 3 | 3 | 8 | 22 | 5 |
| Sarcoma | 16 | 2307 | 34.9%<br>[8.3, 76.2]<br>(I<sup>2</sup>=94.2%,<br>p<0.0001) | 19.2%<br>[9.2, 36.0]<br>(I<sup>2</sup>=95.8%,<br>p<0.0001) | 1.4% [0.9, 2.4]<br>(I<sup>2</sup>=0.0%, p=0.662) | 82.9%<br>[22.7, 95.1]<br>(I<sup>2</sup>=97.9%,<br>p<0.0001) | NA | 56.9% [40.2, 69.1]<br>(I<sup>2</sup>=80.6%,<br>p<0.0001) | 36.0% [30.0, 42.5]<br>(I<sup>2</sup>=81.9%,<br>p<0.0001) | NA |
|  |  | k | 5 | 4 | 3 | 3 |  | 8 | 16 |  |

Data is presented as value with [95% confidence interval]
k, number of included studies for analysis; N, total number of participants; NA, not available.

**Figure 1.**
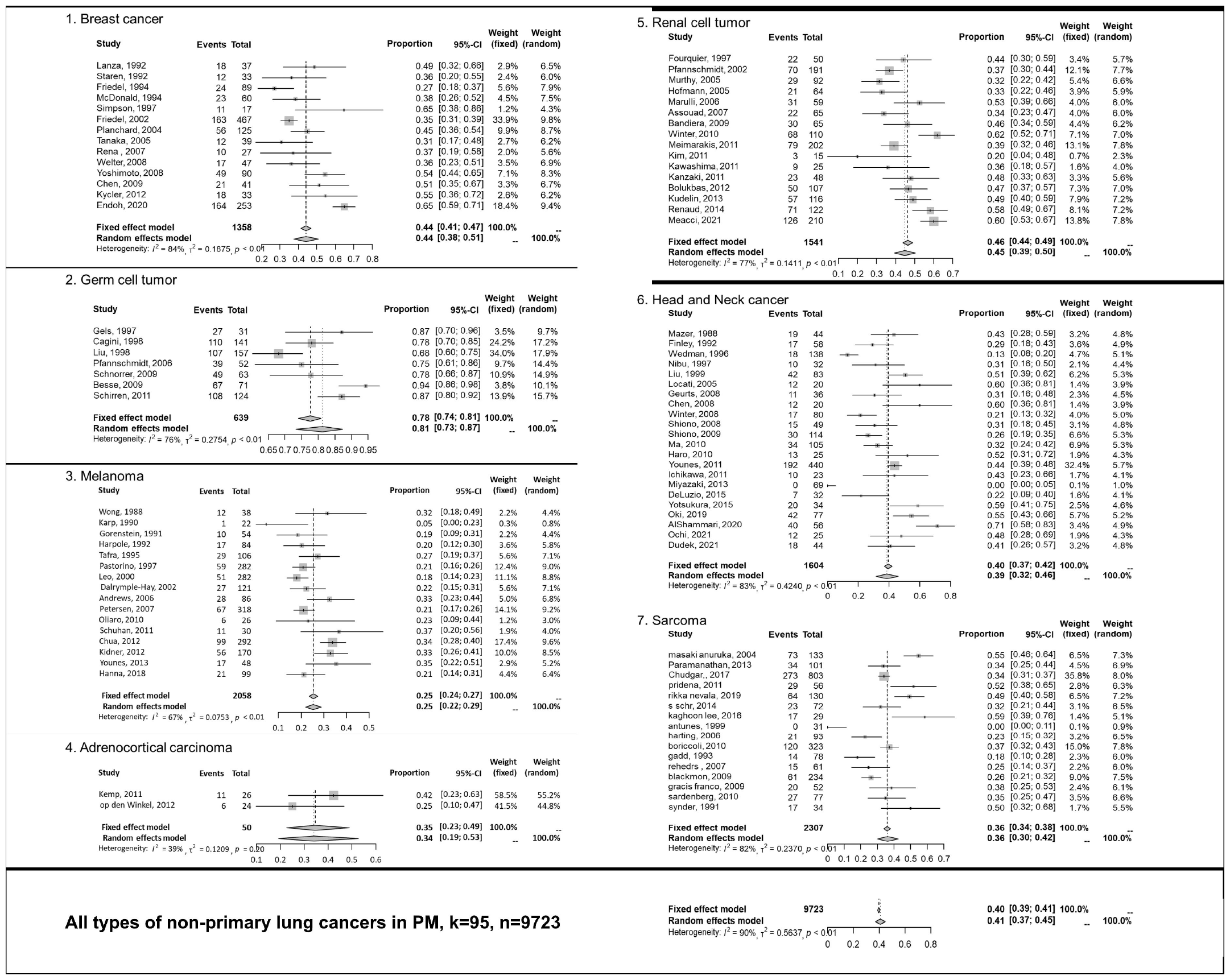
Forest plots of five-year overall survival rate after pulmonary metastasectomy according to primary cancer types

**Figure 2.**
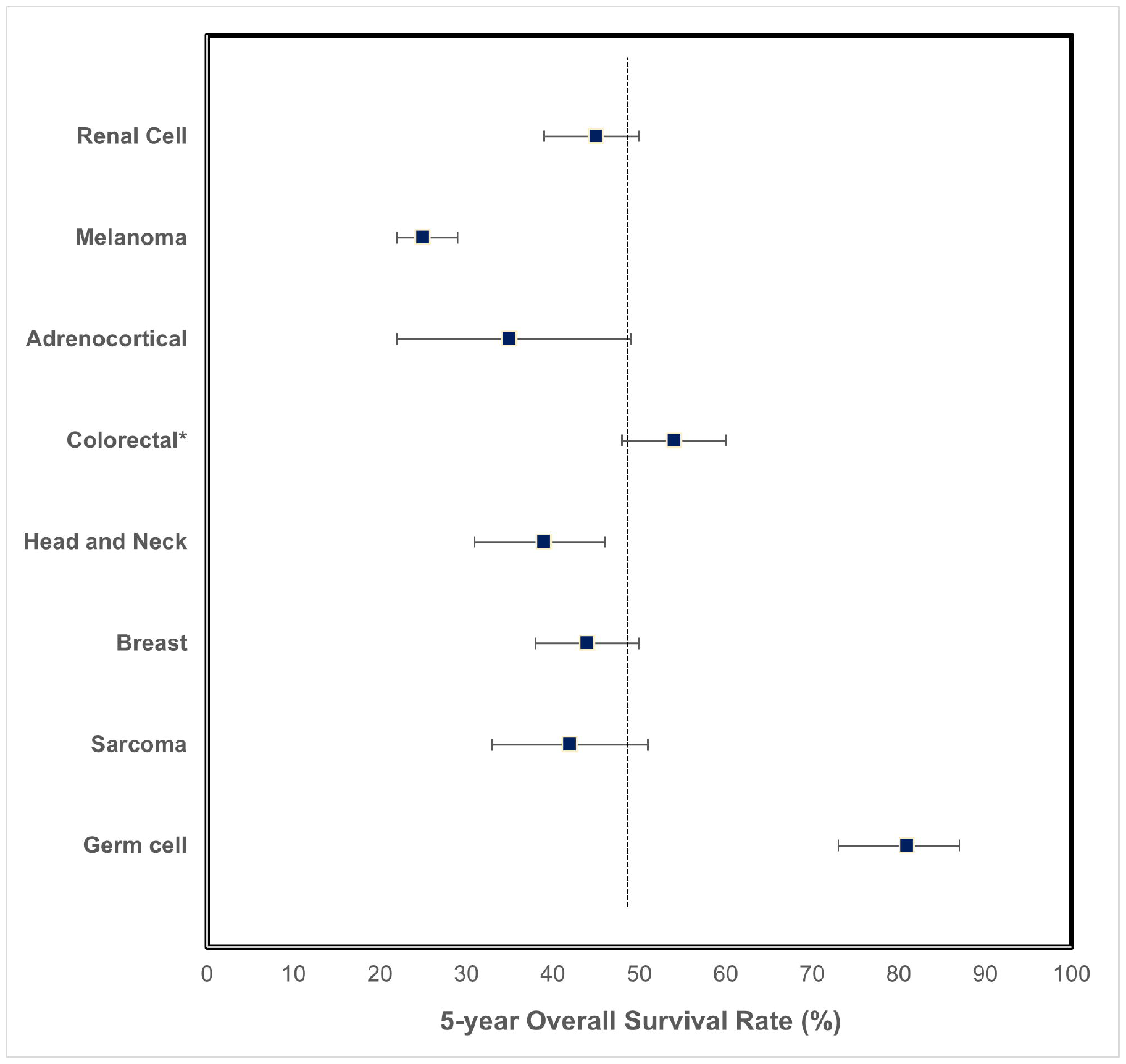
Summarized overall survival according to primary cancer types

## Discussion

This umbrella review provides a comprehensive overview of PM for non-primary lung cancer in terms of patients and tumor characteristics alongside clinical outcomes. Due to the limited number of patients and complex medical history of these patients, there has not been a generalized review of this topic. Clinicians also do not have comprehensive understanding of these patients, often relying on institutional-level evidence. However, this study could facilitate more systematic and objective evaluation of the clinical benefits of PM. To of our knowledge, this is the first attempt to analyze PM outcomes in non-primary lung cancers, which have a relatively low incidence.

The “Seed and Soil” hypothesis, proposed by Stephen Paget in 1889 based on an autopsy analysis, remains as a backbone of biology behind metastasis.^2^ The characteristics of tumor ‘seeds’ were explained by loss of adherence,^25^ epithelial to mesenchymal transition,^26^ increased cell motility,^27^ evasion of apoptosis,^28^ and extravasation.^29^ In addition, the expression of specific genes could promote organ-specific metastasis; breast cancer lung metastasis was related to SIRT7, VCAM-1, and Cx43.^30^ Though the mechanics of lung could be a contributing factor since it is the end organ with extensive capillary blood supply,^2^ successful colonization in the lung can be explained by lung-specific microenvironmental factors. Extrathoracic cancers secreting factors and extracellular vesicles adjust the lung microenvironment to transit into pre-metastatic niches.^31^ Cytoskeletal anchoring proteins such as ezrin^32^ and pathways involving nuclear factor kB^33^ and transforming growth factor ß^34^ have been associated with lung metastasis. Chemokines produced in the lung such as CXCL12 and CXCR4 could also stimulate tumor cells to infiltrate.^35–37^ Nonetheless, further investigations are warranted to delineate the key steps resulting in lung metastasis and find targetable pathways for therapeutic development aimed at minimizing pulmonary metastasis.

Germ cell tumors exhibit significantly better clinical outcomes than other cancers. Other than tumor characteristics, demographic differences might have a pivotal role in these results; patients with germ cell tumors were typically younger than other groups. Thus, medical comorbidity and functional capacity would be much better preserved in this group. Contrary to germ cell tumors, melanoma demonstrates the worst clinical outcomes post-PM, likely due to its ability to evade the host immune response, angiogenic characters, and ‘stemness’ traits of melanocytes.^38^ These negative oncologic characteristics might lead to worse outcomes with pulmonary metastatic melanoma. Other than these two primary cancers, however, most primary cancers had similar long-term clinical outcomes post-PM regardless of their histology, suggesting a pragmatic approach towards conducting pan-tumor prospective or randomized clinical trials to delineate the role of PM in real-world settings.^39^

Apart from primary cancer characteristics, the surgical procedure itself is an essential part in achieving optimal outcomes post-PM. Surgeons evaluate whether those lesions can be completely resectable or not.^8^ In this analysis, the complete resection rate was around 80%. Less than 90-100% of complete resection rate could be due to clinical scenarios that PM is performed, such as tissue confirmation of metastasis or central lesion involvement. Other factors such as extensive pulmonary involvement of metastatic lesions or non-systematic guidance for surgical candidates could have led to this non-optimal result. The criteria for acceptable PM was suggested by Kondo et al.,^40^ but there is no objective criteria to define surgical resectability and mostly depends on surgeons. Since many studies described superior outcomes in complete resection groups compared to incomplete ones, there needs to be more generalized guidance to find suitable candidates for PM. Furthermore, the benefits of extensive surgery, such as pneumonectomy and lung transplant, should be explored as alternative approaches for extensive pulmonary metastasis refractory to other multimodality treatments. A prospective clinical trial is ongoing to evaluate the role of bilateral lung transplants as curative PM and lung function preservation for metastatic cancers to the lung only (NCT 05671887).^41^ Recent advancements in technology and survival of lung transplant could also selectively be applied to some patients.^42,43^

The integration of liquid biopsy in PM has brought new insights into this field. Metastasectomy is related to lower levels of circulating-tumor DNA (ctDNA) and decreased the risk of recurrence. Lee et al. reported that most of the ctDNA was cleared after metastasectomy for metastatic colorectal cancer in their prospective study.^44^ The benefit of lower ctDNA levels during postoperative follow-up has also been suggested during multimodality treatment of locally advanced cancers.^45^ Clinical applications of liquid biopsy in PM could facilitate early recurrence detection and improved survival outcomes since most cancers exhibited high recurrence rates.

Despite the valuable insights provided by this study, there are several limitations in this study that should be acknowledged. First, there was limited data in terms of neoadjuvant or adjuvant chemo/radio/immunotherapy among patients undergoing PM, which restricts our understanding of how these treatment modalities influence outcomes in these patients. Second, the impact of different surgical modalities or demographic factors on clinical outcomes were not accessible from most studies. Distinguishing the survival benefit derived solely from surgery versus outcomes resulting from the natural progression of cancer remains challenging. Due to difficulties, or even impossibility, of having control groups in these issues, the results of PM should be cautiously applied. Third, significant heterogeneity observed across most meta-analyses underscores the need for cautious interpretation of results. Moreover, the absence of prospective studies and the diverse nature of included studies warrant careful consideration of the findings presented.

In conclusion, PM for non-primary lung cancer is still a valuable treatment option, offering fair clinical outcomes for patients in advanced stages. While ongoing clinical trials with medical treatments continue, the 30-40% five-year survival rates observed post-PM highlight its continued relevance and the need to further explore its benefits.

## Supporting information

PRISMA

## Data Availability

All data produced in the present study are available upon reasonable request to the authors

## Acknowledgements

None.

## Data Availability Statement

The data underlying this article will be shared by the corresponding author upon reasonable request.

